# Transitory impact of subclinical *Shigella* infections on biomarkers of environmental enteropathy in children under 2 years

**DOI:** 10.1101/2024.12.18.24319210

**Authors:** Haley A. Liakakos, James A. Platts-Mills, Maria Garcia Quesada, Jie Liu, Eric R. Houpt, Elizabeth T. Rogawski McQuade

## Abstract

Clinical and subclinical *Shigella* infections among children living in low- and middle-income countries (LMICs) have been associated with long-term adverse effects such as impaired linear growth. The mechanism for the impact of subclinical infections has been theorized to occur through contributions to environmental enteropathy (EE). While *Shigella* has previously been associated with biomarkers of EE at the time of infection, we evaluated whether this impact was sustained after infections, which would support EE being the mechanism for the effects of *Shigella* on growth. A prospective birth cohort study of 1,715 children living in 8 different LMICs was conducted. Over the course of 24 months, monthly non-diarrheal stool samples were analyzed for subclinical *Shigella* infections through quantitative PCR methods. EE was reflected by elevated concentrations of 3 fecal biomarkers: myeloperoxidase (MPO), neopterin (NEO), and alpha-1-antitrypsin (AAT). MPO concentrations were found to be significantly higher by 0.30 log(nm/mL) (95% CI: 0.23, 0.37) in the initial month of *Shigella* detection among stools with subclinical *Shigella* infections. After the *Shigella* infection, MPO concentrations declined throughout the following 6 months, and concentrations were lower by 6 months post-infection [MPO 6-month difference: -0.16 log(nm/mL) (95% CI: -0.26, -0.04)]. Subclinical *Shigella* infections had no effect on NEO concentration levels within the initial month of *Shigella* detection but did decrease post-infection. Subclinical *Shigella* infections had no effect on AAT concentration levels until 6 months post-infection [AAT difference: -0.13 log(mg/g) (95% CI: - 0.24, -0.03)]. These findings did not differ by antibiotic use around time of index infection. The impact of *Shigella* on biomarkers of EE was not sustained, suggesting the negative association between *Shigella* and growth could be explained by the accumulation of time-limited rather than persistent effects on inflammation.

## Introduction

*Shigella* is an infectious bacterial pathogen which causes shigellosis, a high burden diarrheal disease that disproportionately affects children under 5 years living in low- and middle-income countries (LMICs) [1–3]. *Shigella* infection is generally thought to be defined by diarrhea and dysentery, but an analysis of the Etiology, Risk Factors, and Interactions of Enteric Infections and Malnutrition and the Consequences for Child Health (MAL-ED) study found that, among children under 2 years, only 15% of *Shigella*-attributed diarrheal stools were accompanied by dysentery and 64% of children that tested positive for *Shigella* never had a *Shigella*-attributed diarrheal episode [4,5]. Subclinical infections, despite having less acute clinical manifestations, have been associated with poor long-term outcomes comparable to clinical infections such as sustained impaired child growth throughout the first 2-5 years of life [6].

While the mechanism for how *Shigella* and other enteric infections could lead to these longer-term adverse effects is unknown, one proposed theory is through environmental enteropathy (EE) [7–9]. EE is a condition of the intestines characterized by inflammation, greater intestinal permeability, and shortening of villi from prolonged pathogen infections which results in malabsorption of nutrients [9,10]. While EE in the short-term is generally asymptomatic, the longer-term impacts have been theorized to include growth stunting, poor cognitive development, and poor vaccine effectiveness in children [9]. *Shigella* infections cause inflammation of the intestines which is theorized to lead to both the clinical symptoms of diarrhea and dysentery and the long-term impact on the intestines through EE [11].

Diagnosing EE is difficult and expensive as it requires invasive intestinal biopsies since physical symptoms can be general and sometimes not apparent [10,12]. Alternatively, there are several fecal biomarkers that capture multiple aspects of gut health that are commonly used in EE analyses [12]. Myeloperoxidase (MPO) reflects inflammation from neutrophil activity. MPO is unique compared to other inflammatory biomarkers as secretion in the stool is not dependent on being breastfed and it provides direct evidence for intestinal disease activity [8,12–14].

Neopterin (NEO) reflects inflammation from TH1 immune activity, and NEO is resistant to being broken down in stool [8,12,13,15]. Alpha-1-antitrypsin (AAT) reflects intestinal permeability, is a clear indicator of protein loss, and is also resistant to being broken down in stool [8,12,13,16,17]. These biomarkers, among others, provide opportunities to non-invasively examine intestinal function and health.

Studies in LMICs have shown consistent findings of above average fecal biomarker levels that have been associated with poor linear growth [7,8,18]. Furthermore, both clinical and subclinical *Shigella* infections have been associated with elevated fecal biomarker concentrations [6–8,11,16,19]. Specifically, *Shigella* has been associated with significant fecal MPO elevations and a dose response relation of MPO increasing by 0.21 logs for every log of *Shigella* quantity detected [4,8]. While the evidence is consistent, one common limitation in these studies stems from the cross-sectional nature of the association between *Shigella* and biomarkers. For *Shigella* to affect longer-term outcomes like growth, we hypothesize the effects of *Shigella* on inflammation and EE would have to persist beyond the initial *Shigella* infection. This study evaluates the longitudinal effects of subclinical *Shigella* infections on fecal biomarkers of enteropathy to interrogate the potential mechanism for *Shigella* to impact growth through EE. Furthermore, even though *Shigella* is often treated with antibiotics, little is known on how antibiotics aid in the recovery of intestines from EE [20]. The second aim of this study is to provide insight about whether the effects of *Shigella* on EE differ depending on recent antibiotic therapy.

## Methods

### Study Population

We performed a secondary analysis of the MAL-ED prospective birth cohort study. From November 2009 through February 2012, infants were enrolled within 17 days of birth and followed for 24 months. There was a focus on resource-limited settings within LMICs, and at least 200 children were enrolled in each of the following 8 sites: Dhaka, Bangladesh; Fortaleza, Brazil; Vellore, India; Bhaktapur, Nepal; Loreto, Peru; Naushahro Feroze, Pakistan; Venda, South Africa; and Haydom, Tanzania. Healthy infants were eligible for enrollment if their caregiver reported they planned to live within the study area for the next 6 months and approved of twice weekly home visits for 24 months. Infants were excluded from study enrollment if they were hospitalized for anything besides a healthy delivery, were diagnosed by a medical doctor for a severe or chronic condition, diagnosed for enteropathies, or weighed less than 1,500 grams at birth. Infants were also excluded if their family anticipated living outside the study area for more than 30 consecutive days within the first 6 months, if the infant was not a singleton, or if the mother was younger than 16 years of age or had another child enrolled in the study [21].

## Data Collection

Field MAL-ED researchers visited households twice weekly to collect basic health and dietary information as well as to complete disease surveillance measures. Sociodemographic information was collected at study enrollment, and updated every 6 months [21]. After 9 months, information on diet and breastfeeding was collected monthly rather than during the twice weekly visits [22]. Enteric pathogen infections and EE biomarker concentrations were measured in non-diarrheal stool samples collected monthly. To ensure clear separation from diarrhea, non-diarrheal samples were taken at least 3 days before or 3 days after a maternal reported diarrhea episode [23]. These samples went under quantitative polymerase chain reaction (qPCR) as previously described for identification of 29 pathogens, including the following which were identified as the most prevalent pathogens in MAL-ED: adenovirus 40/41, astrovirus, *Campylobacter, Cryptosporidium, Entercytozoon bieneusi*, enteroaggregative *Escherichia coli* (EAEC), enterotoxigenic *Escherichia coli* (ETEC), typical enteropathogenic *Escherichia coli* (tEPEC), atypical enteropathogenic *Escherichia coli* (aEPEC), *Giardia*, norovirus, sapovirus, and *Shigella* [6]. To measure the EE biomarkers, quantitative ELISAs were run for MPO, NEO, and AAT on the non-diarrheal stool samples collected on months 1-12, 15, 18, 21, and 24 [21]. Data on recent antibiotic use of any drug class, cephalosporins, macrolides, and fluoroquinolones were collected during the twice weekly household questionnaires [23].

## Statistical Analysis

*Shigella* infections were defined in non-diarrheal stool samples by qPCR detection of the *ipaH* gene at a quantitative cycle threshold of less than 35. We estimated the effects of *Shigella* infections on each EE biomarker at the time of the index *Shigella* infection through 6-months post-infection using multivariable linear regression. Specifically, for each non-diarrheal stool, we estimated the association of *Shigella* infection compared to no *Shigella* infection with MPO, NEO, and AAT concentrations respectively during the same (index) month and for each following month up to 6-months post-infection in separate models. Each model was adjusted for country of residence, age at the time of the infection, sex, whether the child had been exclusively breastfed up until time of the infection, stool consistency, previous *Shigella* infection, and infections with other enteric pathogens. Recent previous *Shigella* infection was defined as *Shigella* detected in another stool sample from the prior 3 months. Infections with other enteric pathogens (coinfections) at the time of the index *Shigella* infection were included in the models if the pathogen was independently associated with the EE biomarker outcome. To determine which pathogens were independently associated with the EE biomarkers, multivariable linear regression models were run for each pathogen (defined by detection at a cycle threshold <35) against stool concentrations of MPO, NEO, and AAT respectively while adjusting for country site and age. The pathogens that were statistically significantly (p<0.05) associated with the biomarker were included as covariates in the models for that biomarker.

In sensitivity analyses, we detrended biomarker concentrations by age to account for naturally higher concentrations in younger children, additionally adjusted the models for coinfections at the time of the biomarker outcome measurement, and further adjusted the models for *Shigella* detection at the time of the biomarker outcome measurement. Another sensitivity analysis subset to *Shigella* infections within the first 18 months of life only to allow for all included infections to have a complete 6 months of follow-up samples.

To determine if recent antibiotic treatment impacted the effect of *Shigella* on the concentrations of MPO, NEO, and AAT, the same linear regression models were run with an interaction term between *Shigella* infection and antibiotic use 15 days before or after the index *Shigella* infection. The different classes of antibiotics examined were cephalosporins, macrolides, and fluoroquinolones, and the last set of models looked at any antibiotic class used.

## Ethics Statement

The MAL-ED study collected data on and from human participants under the age of 2 along with parental consent. The study design was approved by the University of Virginia’s institutional review board. Approval was given within each of the 8 sites. The Bangladesh site had approval from the Ethical Review Committee, International Centre for Diarrhoeal Disease Research, Bangladesh. The Brazil site had approval from the Committee for Ethics in Research, Universidade Federal do Ceara, and the National Ethical Research Committee, Health Ministry, Council of National Health. The India site had approval from the Institutional Review Board, Christian Medical College, Vellore, and the Health Ministry Screening Committee, Indian Council of Medical Research. The Nepal site had approval from the Institutional Review Board, Institute of Medicine, Tribhuvan University, the Ethical Review Board, Nepal Health Research Council, and the Institutional Review Board, Walter Reed Army Institute of Research. The Peru site had approval from the Institutional Review Board, Johns Hopkins University, and the PRISMA Ethics Committee, Health Ministry, Loreto. The Pakistan site had approval from the Ethical Review Committee, Aga Khan University. The South African site had approval from the Health, Safety and Research Ethics Committee, University of Venda, and the Department of Health and Social Development, Limpopo Provincial Government. The Tanzania site had approval from the Medical Research Coordinating Committee, National Institute for Medical Research, and the Chief Medical Officer, Ministry of Health and Social Welfare. This study’s secondary analysis did not require approval from Emory University’s institutional review board, as it did not fall under human subjects research since the data from the primary MAL-ED study is public and deidentified.

## Results

### EE Biomarker Concentrations Through 6 Months After Shigella Infection

Over the course of 2 years, 1,715 children in the MAL-ED study contributed 34,654 non-diarrheal stool samples. Of these, 10% (N = 3,505 samples) tested positive for *Shigella*. Details of demographic characteristics associated with stool samples with and without *Shigella* detected are in Table 1. In stools with subclinical *Shigella* infections, the inflammatory EE biomarker, MPO, was elevated by 0.30 log(ng/mL) (95% CI: 0.23, 0.37) compared to stools without *Shigella* during the initial month of *Shigella* detection (Fig 1; S1 Table). In the stools 1-4 months following the identification of the subclinical infection, the concentration of MPO was no different compared to those following stools without *Shigella*. After 5-6 months, MPO concentration levels were 0.18 log(ng/mL) (95% CI: 0.00, 0.35) and 0.16 log(ng/mL) (95% CI: 0.04, 0.26) lower following a *Shigella* infection compared to samples following no *Shigella* infection. There was no clear difference in NEO concentrations, the EE biomarker characterizing gut immunity, in the initial detection month between those with and without subclinical *Shigella* infections [NEO difference: 0.01 log(nmol/L) (95% CI: -0.05, 0.07)]. The subsequent 6 months showed that stools following subclinical *Shigella* infections had lower concentrations of NEO compared to those not following an infection (Fig 1; S1 Table). Similarly, in the initial month of detecting subclinical *Shigella* infections, there was no clear difference in concentrations of the EE biomarker AAT, reflecting intestinal permeability [AAT difference: -0.02 log(mg/g) (95% CI: -0.08, 0.04)]. AAT concentrations were lower among stools following a subclinical *Shigella* infection, particularly after 6 months [AAT difference: -0.13 log(mg/g) (95% CI: -0.24, -0.03)], compared to stools 6 months after no *Shigella* infection (Fig 1; S1 Table).

**Table 1.**
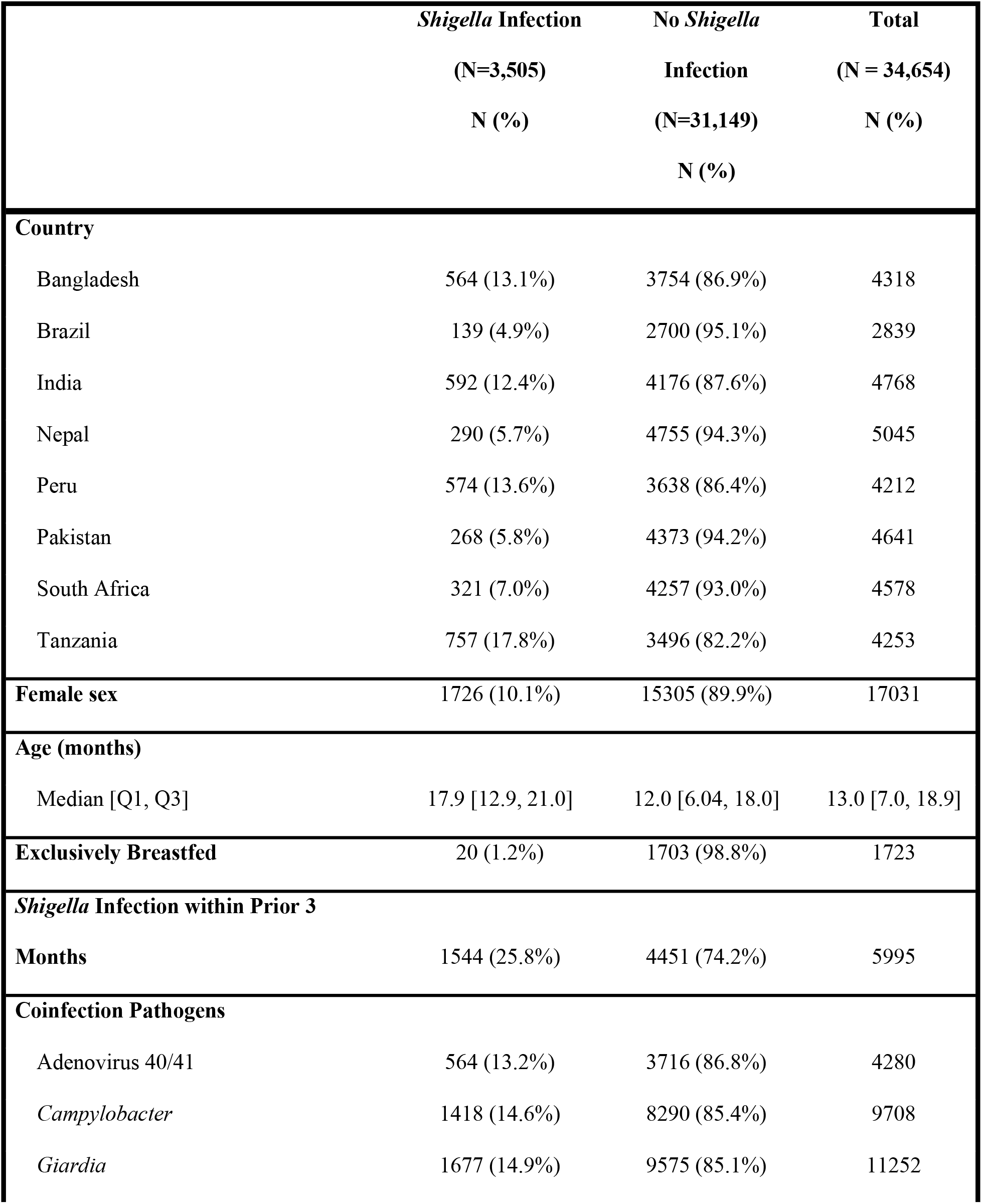
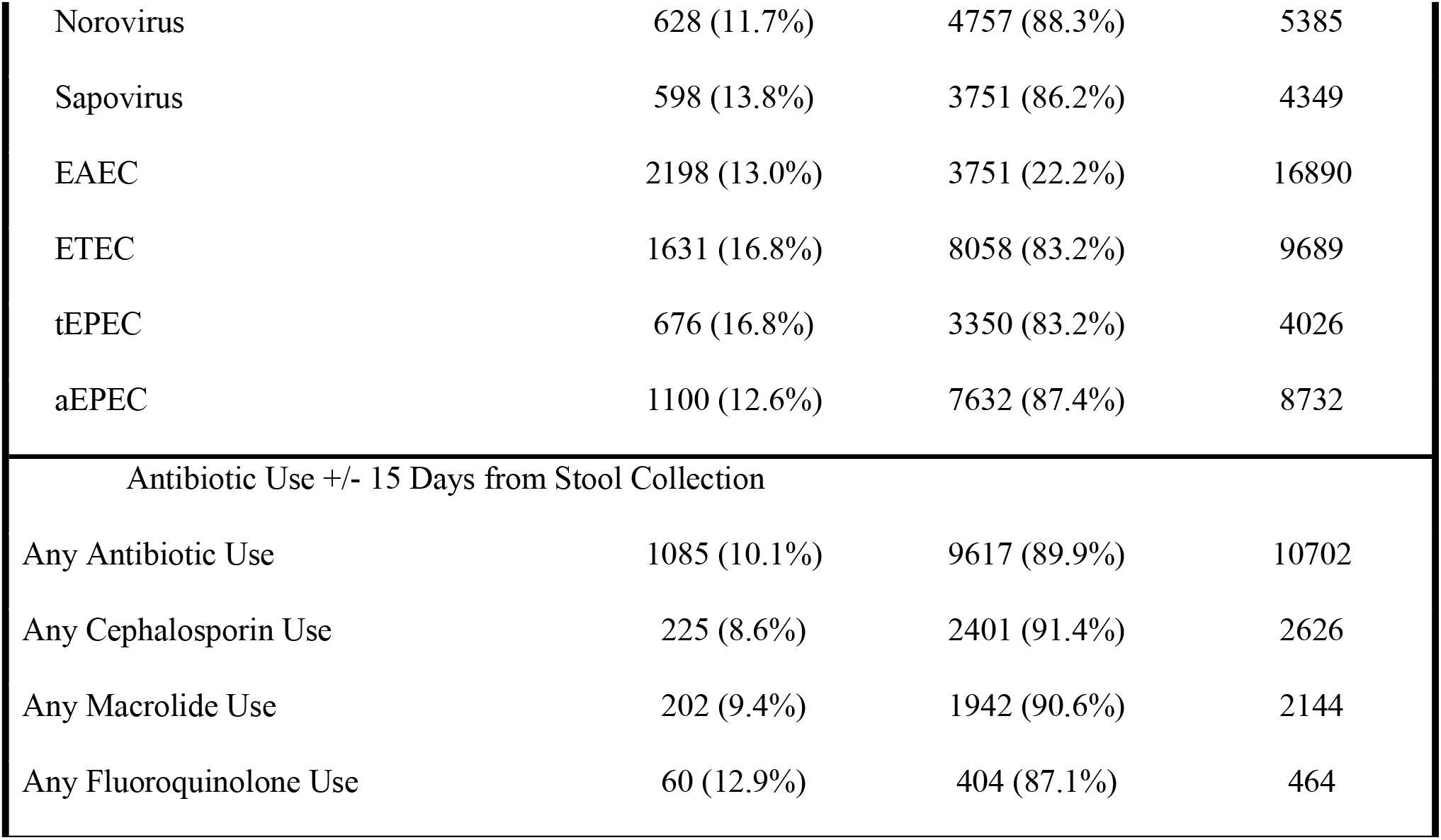
Characteristics of non-diarrheal stool samples from 1715 children in MAL-ED study.

**Fig 1.**
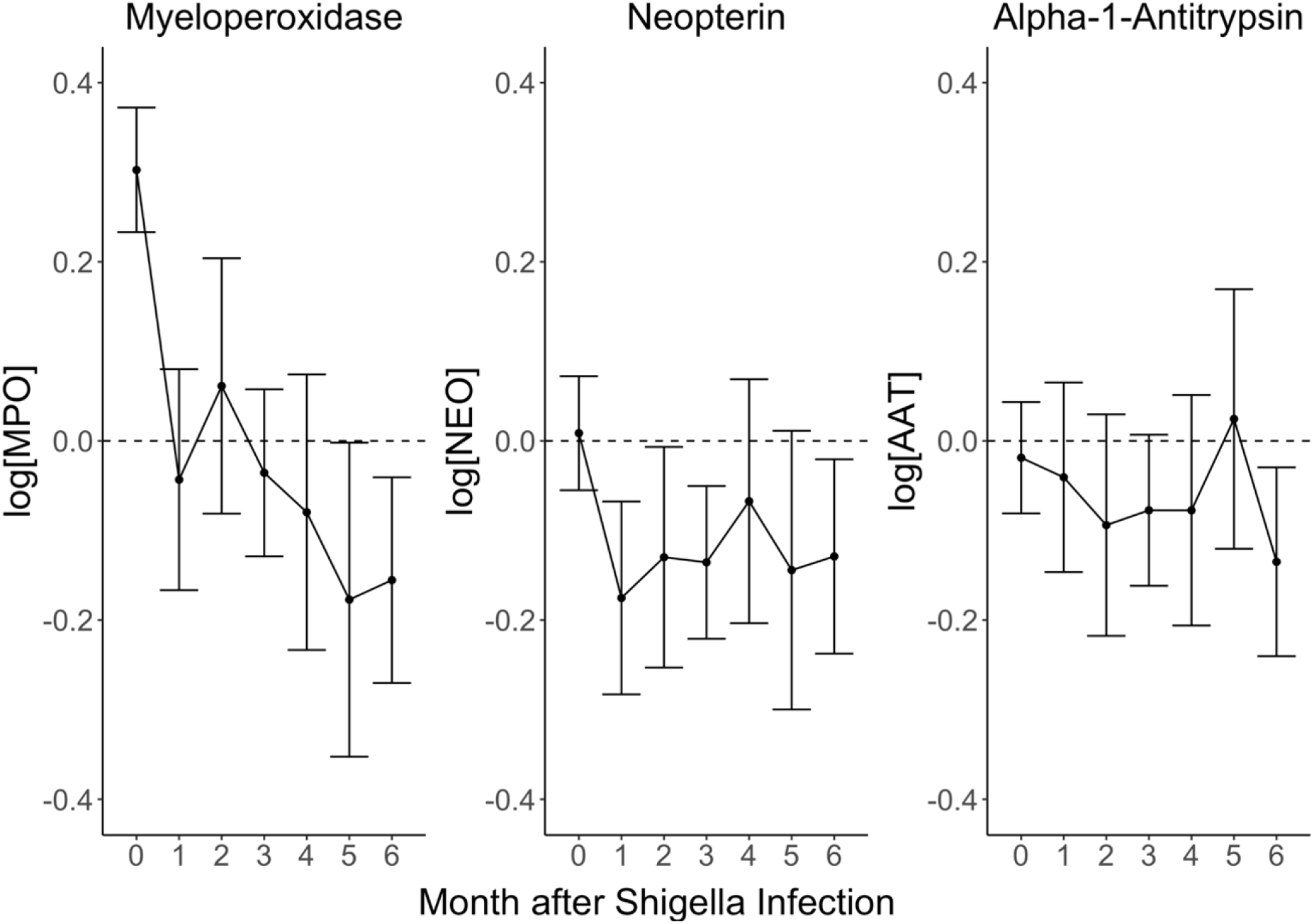
Association of *Shigella* infections with EE biomarker concentration over time. Each plot shows EE biomarker log concentration differences and 95% confidence intervals comparing non-diarrheal stool samples with and without *Shigella* detection at month 0. Concentrations were measured monthly from the index *Shigella*-positive detection through 6-months post-detection.

In sensitivity analyses, the associations between *Shigella* infection and MPO biomarker concentrations showed little difference when adjusting for age, alternatively when detrending the biomarker concentrations for age, or by doing both. The results were also very similar when additionally adjusting for other infections at the time of the biomarker measurement, including subsequent *Shigella* infections (S1 Fig). Finally, results were similar when restricting the analysis to stools collected at 18 months of age or younger (S2 Fig).

### Modification by Antibiotic Treatment

There were no consistent differences in the impact of *Shigella* on MPO, NEO, or AAT concentrations between children who were and were not treated with a cephalosporin, macrolide, fluoroquinolone, or any antibiotic in general with a 15-day range of the index *Shigella* infection (Fig 2). Children who took macrolides within the 15 days before or after the *Shigella* infection had lower MPO concentrations starting around 4 months after the infection compared to children without a *Shigella* infection who took macrolides. Specifically, MPO was significantly lower with *Shigella* among children with macrolide antibiotic use 15 days before or after infection at 5-months post-infection only. In contrast, while estimates were often imprecise, NEO concentrations were more elevated after *Shigella* infections when antibiotics were used. The impact of antibiotic use on *Shigella’s* impact on AAT concentrations was inconsistent over time.

**Fig 2.**
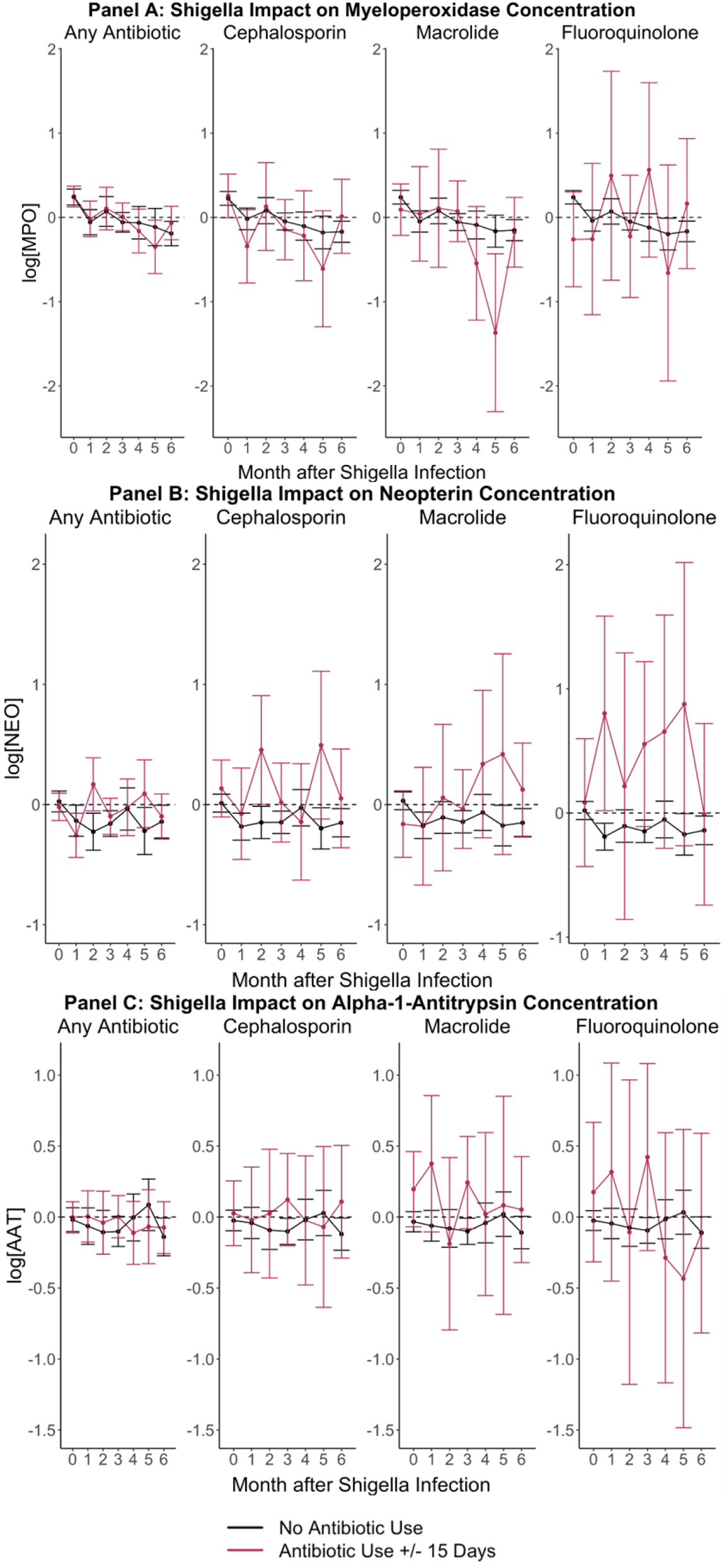
Associations between *Shigella* infections and EE biomarker concentrations from time of index subclinical *Shigella* detection through 6-months post-detection, stratified by antibiotic use within the 15 days before or after *Shigella* infection. Each panel shows the respective EE biomarker log concentration differences comparing non-diarrheal stool samples with and without *Shigella* detection at month 0. Concentrations were measured monthly from the index *Shigella*-positive detection through 6-months post-detection. The black line represents no antibiotic use, and the red line represents antibiotic use within the 15-days before or after month 0.

## Discussion

In the MAL-ED study, subclinical *Shigella* infections were associated with elevated MPO concentrations at the time of index infection, but concentrations returned to baseline by one month after the infection, showing no evidence of sustained impact on inflammation. Subclinical *Shigella* infections were not associated with elevated NEO or AAT concentrations within the initial month of infection nor the following 6 months. By 6-months post-infection, all 3 biomarker concentrations tended to be lower following stools with an index *Shigella* infection compared to those following no index infection. These associations were not explained by subsequent infections. There was limited and inconsistent evidence that antibiotic use around time of infection modified the impact of *Shigella*.

Elevated concentrations of MPO, NEO, and AAT have been associated with risk for growth stunting among young children [12]. Children naturally have higher levels of these biomarkers compared to adults, with concentrations decreasing with age. Even so, compared to high-income countries, children living in LMICs have further elevated levels [8,18]. The observed effects of *Shigella* infection on EE fecal biomarkers during the initial month of infection were consistent with prior research: MPO concentrations were elevated at time of infection and there was not a clear difference in NEO and AAT concentrations compared to those not infected [4,8,19]. However, the fact that MPO was only transiently elevated at the time of the index infection indicates that MPO is unlikely to be a good marker of the sustained impact of *Shigella* on EE and growth. In contrast, the kinetics of MPO following *Shigella* infections could make it a valuable marker of invasive bacterial enteric infection when children present to care [26].

Previous studies have found that antibiotic treatment can mitigate the impact of diarrhea, and specifically *Shigella* diarrhea, on growth [24,25]. Our findings provide evidence that the biomarker concentrations differ only slightly, if at all, between *Shigella* infections treated with fluoroquinolones, cephalosporins, and macrolides and those that were untreated. Because MPO concentrations returned to baseline one month following the index infection, we were unable to observe shorter term effects, for example whether antibiotics could hasten recovery from inflammation within the first weeks following infection.

The MAL-ED study design allowed for the unique longitudinal analysis of the effect of subclinical *Shigella* infections on EE fecal biomarker concentrations among children across 8 global sites, while accounting for age, breastmilk consumption, and other enteric infections which have been associated with inflammatory biomarkers [18]. This study was limited by not all non-diarrheal stool samples being tested for biomarker concentrations. Monthly samples in the second year of life were tested quarterly for biomarker levels which gave the infections in the first year of life a heavier weight in the analysis. Furthermore, because the biomarkers were not measured in diarrheal stools, we were unable to examine the potential sustained impact of *Shigella* diarrhea. Finally, there is lack of literature on healthy biomarker levels among children in both high-income countries and LMICs. While the levels between those with and without *Shigella* infections can be compared, it is not clear which concentration levels contribute to EE and/or growth impairment.

Many studies have investigated which enteric pathogens contribute to EE, and *Shigella* has consistently been found to be a high burden pathogen in LMICs with long-term effects on EE related to both symptomatic and asymptomatic infections [1,3]. This study concludes that if *Shigella’s* long-term effects are brought upon through a sustained impact on EE, these effects are not captured by the biomarkers studied here. Stronger associations between enteric pathogens and EE biomarkers may arise when multiple pathogens are considered given many children in these regions experience co-infections, which may have a bigger impact on the long-term health of those infected. A better understanding of how *Shigella* affects EE and of the mechanisms for the long-term effects of *Shigella* on child growth and development is needed.

## Data Availability

The data underlying the results presented in the study are available from the ClinEpiDB database (https://clinepidb.org/ce/app/workspace/analyses/DS_5c41b87221/new/details). One must register and request access to download data data can be downloaded after a committee reviews the request and grants access.

## Acknowledgement

This work was supported by the National Institutes of Health, National Institute of Allergy and Infectious Diseases (R01AI185140 to ETRM). The Etiology, Risk Factors and Interactions of Enteric Infections and Malnutrition and the Consequences for Child Health and Development Project (MAL-ED) was a collaborative project supported by the Bill & Melinda Gates Foundation (OPP1131125), the Foundation for the NIH, the National Institutes of Health, and the Fogarty International Center.

## Authors’ contributions

HAL led data analysis, visualization, interpretation, and writing of the report. JAP-M and MGQ contributed to data analysis and methodology. JL led the development of the laboratory assays.

ERH led funding acquisition and administration of the parent study. JAP-M, MGQ, JL, and ERH contributed to reviewing/editing the report. ETRM led conceptualization, methodology, and funding acquisition, and contributed to writing of the report.

